# Assessment Methods of Methamphetamine Use in HIV-Positive Populations: A Review of the Literature

**DOI:** 10.1101/2025.09.03.25334488

**Authors:** Margaret H. White

## Abstract

**Background:** Methamphetamine (MA) use is disproportionately higher among people living with HIV (PWH) and contributes to adverse clinical outcomes. Accurate measurement of MA use is essential for research and care but remains inconsistent.

**Methods:** A systematic review of PubMed, Embase, and CINAHL identified 22 peer-reviewed studies (2019–2024) assessing MA use in HIV-positive adults. Data were extracted on measurement methods, demographic composition, and HIV outcomes.

**Results:** Studies employed self-report, urine or hair toxicology, and medical record review. Reporting varied widely in timeframe, frequency, and specificity, with limited detail on route of administration. Most studies were U.S.-based, disproportionately male, and rarely included women or gender-diverse participants. Inconsistent definitions and reliance on broad stimulant categories limited comparability.

**Conclusions:** Current methods for assessing MA use in PWH lack standardization. Greater consistency, inclusion of biological validation, and expanded demographic representation are needed to strengthen research and clinical interventions.

---

Currently, 39 million people are living with HIV (PLWH) globally, with an estimated 1.2 million in the United States (Centers for Disease Control and Prevention [CDC], 2020.) Advances in Antiretroviral therapy (ART) have significantly reduced mortality rates and opportunistic infections (Bradford et al., 2007). Despite these advancements, only 66% of individuals in the United States with HIV are virologically suppressed (CDC, 2020).

Methamphetamine is a synthetic stimulant used recreationally that has a high potential for abuse (Bryson, 2015). From 2015 to 2019, Methamphetamine usage in the United States increased by 43% (Han et al., 2021). methamphetamine use disproportionately affects PLWH, with a rate approximately 10 times higher than the general population (Jones et al., 2022). People with HIV who use methamphetamine are less likely to achieve viral suppression and are less engaged in their HIV primary care (Carrico et al., 2019; Devos et al., 2023; Traeger et al., 2012).

Understanding the scope of methamphetamine use in this group is essential to direct public health strategies and clinical interventions. Accurately capturing methamphetamine use presents methodological challenges due to variability in reporting methods, differences in screening tools, and inconsistent definitions of methamphetamine use across studies. This heterogeneity creates difficulties in synthesizing findings.

The research to date has not produced any reviews assessing the methodology of methamphetamine use. Previous reviews have looked independently at behavioral practices related to methamphetamine use in this population (Maxwell et al., 2019; Moradi et al., 2022), epidemiologic profiles of methamphetamine use (Lodge et al., 2024), or targeted interventions (Rajasingham et al., 2012). The purpose of this review is to synthesize the available data on methodologies used to assess methamphetamine use in adults living with HIV in clinical research. It will explore the question: In adults living with HIV, what methods are used to report and assess methamphetamine usage in research?

## Background and Significance

Methamphetamine usage in the United States has been increasing: among individuals seeking treatment for opioid use disorder (OUD), methamphetamine use rose by 85% from 2011 to 2018 (Cicero et al., 2020). Between 1999 and 2021, methamphetamine-related overdose fatalities surged 50-fold, partly due to the rise in fentanyl adulteration (Han et al., 2021). The psychological impacts of methamphetamine foster dependency and risky behaviors.

Methamphetamine use alone is linked to considerable morbidity and mortality (Han et al., 2021), posing a significant public health challenge. The death rate involving psychostimulants, including methamphetamine, has increased by 317% from 2013 to 2019 (Mattson et al., 2021).

Methamphetamine use is more common in specific demographic groups, including younger individuals, racial minorities, and those with lower socioeconomic status, who are at greater risk for housing instability and lack of insurance (Han et al., 2021).

Methamphetamine use (PLWH) is linked to adverse clinical outcomes, driven by its effects on the immune system and influenced by social determinants of health. Chronic use correlates with adverse health outcomes that can hinder HIV management: increased absenteeism from HIV care, a higher incidence of major depressive disorder, and elevated rates of hepatitis C, syphilis, chlamydia, and gonorrhea (Bamford et al., 2022). These individuals are also less likely to achieve viral suppression and more prone to CD4+ helper T cell counts below 200 cells/mm^3^ (Ross et al., 2023), a sign of immunosuppression and susceptibility to opportunistic infections. Methamphetamine use in the setting of HIV exacerbates disease progression, resulting in adverse health outcomes, including heightened rates of cardiovascular diseases and neurocognitive deterioration. (Carrico et al., 2019).

Accurately measuring and recording methamphetamine use presents several challenges. Methamphetamine (MA) use can be documented in various ways depending on the context of use and the method of administration. Methamphetamine can be identified own its own, or fall under the categories of amphetamine use, stimulant use, or injection drug use. Amphetamines can include other illicit amphetamines as well as prescription amphetamines such as dextroamphetamine, commonly prescribed for attention-deficit hyperactivity disorder. (ADHD). Stimulants can include non-amphetamine substances such as cocaine and even caffeine. While methamphetamine can be injected intravenously, it can also be smoked, snorted, or inserted rectally, so the categorization solely as a drug for injection use can be misleading. Each route can present different health risks, including higher rates of HIV transmission among injection drug users (Bamford et al., 2022). There is no single generally accepted instrument to evaluate methamphetamine use. The National Institute on Drug Abuse (NIDA) cites five screening tools for substance and alcohol use. Only one, the Tobacco, Alcohol, Prescription medication, and other Substance use Tool (TAPS), collects specific information about methamphetamine use (NIDA, 2023). The TAPS asks about current or recent use of substances such as “cocaine, crack, or methamphetamine (crystal meth)” (McNeely et al., 2016) but does not collect detailed information beyond this level of specificity. The National Survey on Drug Use and Health (NSDUH) is a federally funded longitudinal study that attempts to collect substance use in the general population (Center for Behavioral Health Statistics and Quality, 2023). Sections in the NSDUH questionnaire include methamphetamine under the special drugs category, consisting of “methamphetamine, cocaine, heroin with a needle, and sniffing of heroin.”

Retrospective reviews of medical charts and data contain ICD-10 diagnosis codes that can potentially identify methamphetamine use. However, methamphetamine use is reported under the category F15 as stimulant use, which does not differentiate methamphetamine from other stimulants, including caffeine (World Health Organization [WHO], 2019). Additionally, the use of diagnosis codes as a criterion can lead to underreporting due to stigma or incomplete documentation. Medical data can also include information from urine drug screens (UDS) or toxicology results to identify methamphetamine use, a measure of objective data. However, these tests are limited by the short half-life of methamphetamine, typically only detecting recent use within 1-3 days (Oyler et al., 2002). The challenge remains in standardizing these methods to ensure consistent and reliable reporting. The complexities and implications of methamphetamine use necessitate a reliable and valid methodology to capture the scope of usage in this population. Methamphetamine hydrochloride is approved by the Food and Drug Administration (FDA) for ADHD and obesity under the trade name Desoxyn as a schedule II drug (Recordati Rare Diseases, Inc., 2015). Desoxyn has a black box warning for high potential abuse and the prescribing guidelines state it should be “prescribed or dispensed sparingly” (Recordati Rare Diseases Inc., 2015). Because of the risks and infrequency in prescribing, it is an almost entirely illicit in the United States. Methamphetamine is studied across a wide range of disciplines including in addiction medicine, psychiatry, epidemiology, and infectious disease, each of which may prioritize different aspects of use in their reporting. This review will highlight their strengths and limitations by critically evaluating the tools and approaches found in the literature. Identifying current methods for capturing methamphetamine use can help inform the development of standardized approaches.

### Theoretical Framework

The syndemics model of health is a framework that examines how multiple health conditions interact within a population, especially when compounded by social, environmental, and structural factors (Singer et al., 2017). This model assumes that co-occurring diseases, or "syndemics," are not only connected biologically but also can be intensified by social determinants of health such as poverty, stigma, and discrimination. The syndemics model of health is a framework that examines how multiple health conditions interact within a population, especially when compounded by social, environmental, and structural factors (Singer et al., 2017). Syndemic frameworks were initially developed to examine the relationship between substance abuse, violence and AIDS-related outcomes (Singer, 1996). This review utilized a modified version of the syndemic coupling framework developed by Peprah et al. (2022) as a theoretical framework (Figure 1). The model conceptualizes methamphetamine use and HIV as interacting systems that form a coupled syndemic, where a disease system (HIV) and a non-disease system (methamphetamine use) interact bidirectionally to influence health outcomes.

**Figure 1.**
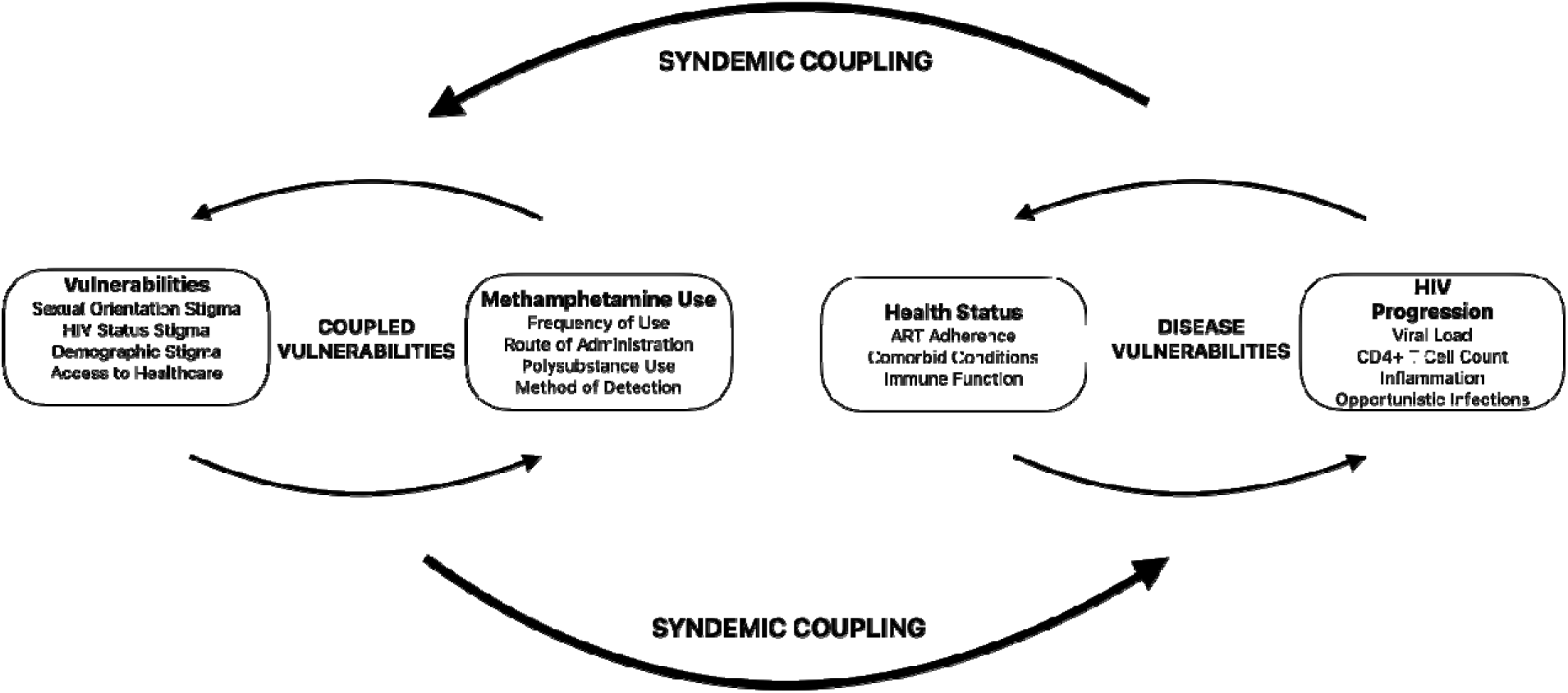
Modified Coupled Syndemic System Conceptual Framework. *Note.* Adapted from "Using a Syndemics Framework to Understand How Substance Use Contributes to Morbidity and Mortality among People Living with HIV in Africa: A Call to Action" by E. Peprah et al., 2022, International Journal of Environmental Research and Public Health, 19(3), p. 7 (https://doi.org/10.3390/ijerph19031097).

**Figure 2.**
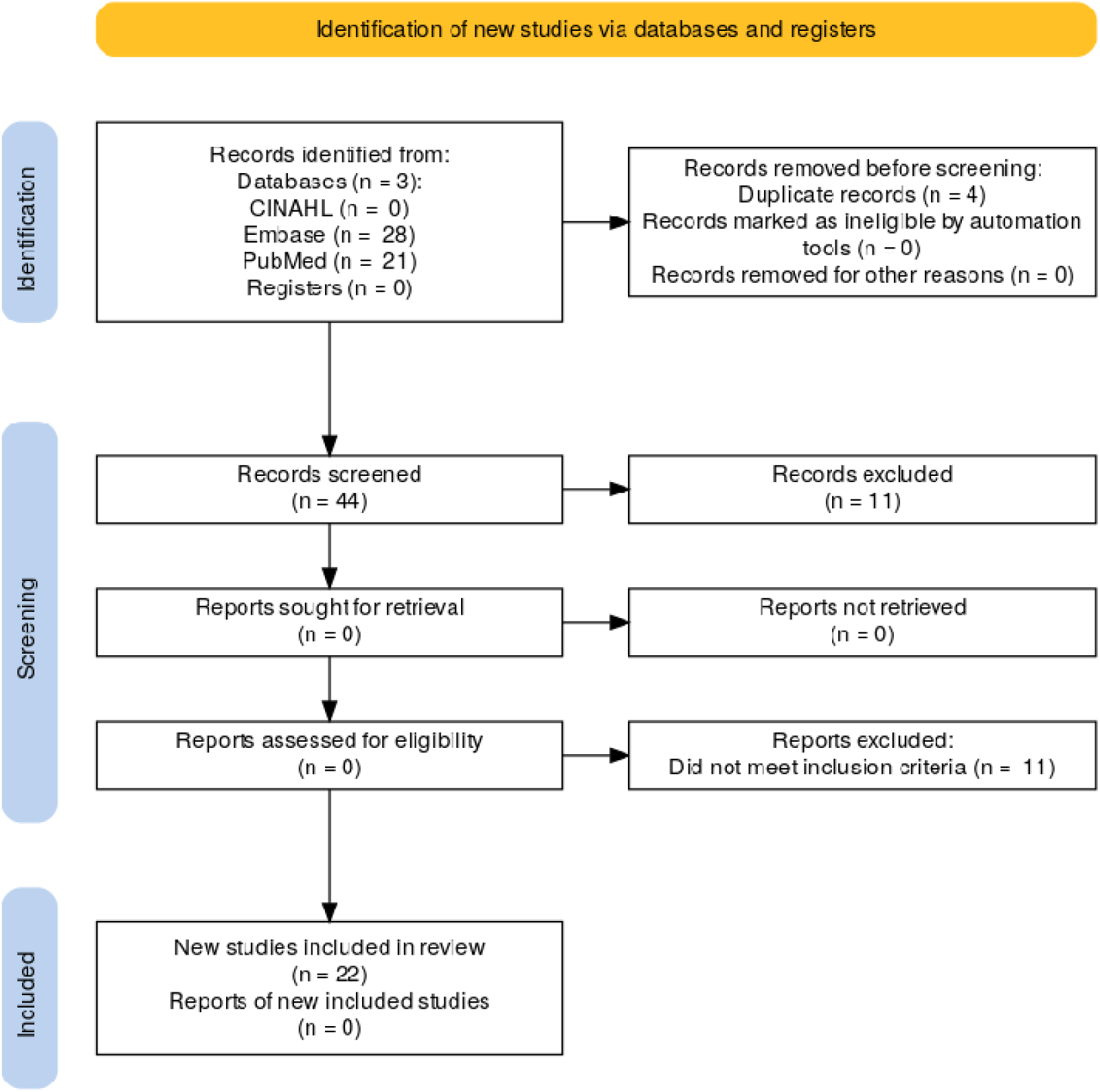
PRISMA Diagram. Note. Adapted from "The PRISMA 2020 Statement: An Updated Guideline for Reporting Systematic Reviews," by M. J. Page, J. E. McKenzie, P. M. Bossuyt, et al., 2021, *BMJ, 372*, n71 (https://doi.org/10.1136/bmj.n71). Copyright 2021 by the authors.

The HIV disease system constitutes the biophysiological (disease) domain. HIV disease activates inflammatory markers, which affect immune function. Methamphetamine use constitutes the nonbiopshysiological (non-disease) domain. Risk behaviors that the environment impacts methamphetamine use can mediate. Methamphetamine use can in turn influence ART adherence as well as impact inflammation and immune function. Concepts in the biophysiological domain can include the HIV viral load, CD4+ T helper cell count, ART regimen, comorbidities, and inflammatory markers. The non-disease system comprises methamphetamine use patterns and associated vulnerabilities, including comorbid substance use, gender and sexual identity, and ART adherence. Under this framework, both biophysiological and nonbiophysiological domains were considered for investigation in the review.

## Methods

### Study Selection and Screening

A comprehensive literature search was conducted to identify empirical studies reporting the methodologies used to document methamphetamine use among adults (age 18 and above) living with HIV. The search focused on how methamphetamine use is operationalized in research, capturing methods such as self-report, biological assays, and medical record documentation. The search followed Preferred Reporting Items for Systematic Reviews and Meta-Analyses (PRISMA) (Page et al., 2021) guidelines. A systematic search of electronic databases using a combination of controlled vocabulary (e.g., MeSH terms in PubMed and Emtree in Embase) and free-text keywords will be used. The search process will include title and abstract screening, followed by full-text review for final eligibility. All search results and reasons for exclusions will be documented using the Covidence platform.

The search was conducted in three key databases—PubMed, Embase, and CINAHL— in assistance with a medical librarian from the Texas Medical Center Library. These databases were selected to represent a multidisciplinary coverage of the literature. PubMed was used to capture peer-reviewed biomedical and clinical research, including randomized controlled trials (RCTs), observational studies, and cohort studies. Embase was selected for its extensive biomedical and psychosocial research, with a focus on studies related to substance use and HIV treatment outcomes. CINAHL was utilized for its literature on healthcare services and nursing research.

The search employed a controlled vocabulary (e.g., MeSH in PubMed and Emtree in Embase) and free-text keywords tailored to each database’s indexing system (Appendix A). The terms were selected to address the three primary constructs: (1) HIV status, (2) methamphetamine use, and (3) methodology or data collection techniques. Boolean operators (AND, OR, NOT) were be used to combine the search terms, optimizing retrieval of relevant studies. Importantly, terms related to pre-exposure prophylaxis (PrEP) were included with the Boolean operator NOT to exclude studies involving HIV-negative populations. The search was restricted to articles published in English between 2019 and 2024 Inclusion criteria for studies consisted of 1) inclusion of HIV-positive participants aged 18 or older, 2) reporting on substance use, 3) English language, 4) published in the last five years as of October 15, 2024. Exclusion criteria were for 1) studies involving non-human subjects 2) populations that fall outside the target age range, 3) studies without explicit mention of methamphetamine use, and 4) studies that focus exclusively on HIV-negative populations or PrEP. Articles were only eligible if they were from peer-reviewed journals. Reviews and pre- prints were not accepted. This population was not limited to men who have sex with men (MSM) to try to ascertain what role methamphetamine use is examined in a female and heterosexual male population.

The initial database search yielded 49 articles. After screening, removal of duplicates, outside peer evaluation and full-text review, 22 articles were selected. Appendix A displays the PRISMA diagram for the literature search. After all articles were selected, data extraction was performed using a codebook (Appendix B). The codebook entries were selected to capture a large amount of variables from the articles for further evaluation for inclusion in the final tables, given the broad nature of the question behind this review.

### Quality Appraisal

The Mixed Methods Appraisal Tool (MMAT) (Hong et al., 2018) was used to evaluate the quality of the included studies. This tool allows for assessing quantitative, qualitative, and mixed-methods studies. The MMAT evaluates studies based on their methodological soundness, sample size, and study design. Only high- and medium-quality studies were included in the final synthesis, while studies with significant methodological flaws or high risk of bias were excluded. Figure 1 summarizes the articles by MMAT category. Although the MMAT does not advocate for a numerical score, articles were categorized by category. All articles included in this review fell under the categories of “moderate-high” or “high” and demonstrated quality.

## Findings

### Characteristics of Included Studies

The study overview and demographic information is provided in Table 2. Of the 22 articles that met the inclusion criteria, four authors contributed multiple studies as lead authors. Carrico et al. authored two articles (2019, 2023), each with different secondary authors, as did Crane et al. (2021, 2022) and Feelemyer et al. (2020, 2024). Lee et al. contributed three studies in 2020 (2020, 2021a, 2021b) with varying co-authorship. Two journals contributed multiple articles: *Drug and Alcohol Dependence* published studies by Crane et al. (2022), Glasner et al. (2022), and Goodman-Meza et al. (2019); the *Journal of Neurovirology* contributed studies by Lee et al. (2020, 2021a), and Kondur et al. (2022). Ten of the studies were secondary data analyses or substudies of larger studies (Carrico et al., 2023; Chahine et al., 2021; Crane et al., 2021; Crane et al, 2022; Feelemyer et al., 2024; Ghanooni et al., 2022; Lee et al., 2020; Lee et al., 2021a; Lee et al., 2021b; Miller et al., 2020), and the study conducted by Carrico et al. (2019) served as the primary study for nine of the included articles (Carrico et al., 2023; Chahine et al, 2021; Ghanooni et al., 2022; Lee et al., 2020; Lee et al., 2021a; Lee et al., 2021b; Miller et al., 2020; Vincent et al., 2021). The majority of the studies were conducted in North America, with seventeen studies from the United States or Canada (Berlin et al., 2024; Carrico et al., 2019, 2024; Chahine et al., 2021; Crane et al., 2021, 2022; Ghanooni et al., 2022; Glasner et al., 2022; Goodman-Meza et al., 2019; Kondur et al., 2022; Korthuis et al., 2021; J. Lee et al., 2020; Lee et al., 2021a; Lee et al., 2021b; Martin et al., 2020; Miller et al., 2020; Sharp et al., 2024; Vincent et al., 2021). The remaining studies were conducted in Asia: three in Vietnam (Feelemyer et al., 2020; Feelemyer et al., 2024; Korthuis et al., 2021), one in Japan (Hayashi et al., 2023), and one in Taiwan (Chen et al., 2021). There were no studies from Europe, Central Asia, Latin America, or Africa.

**Table 1.**
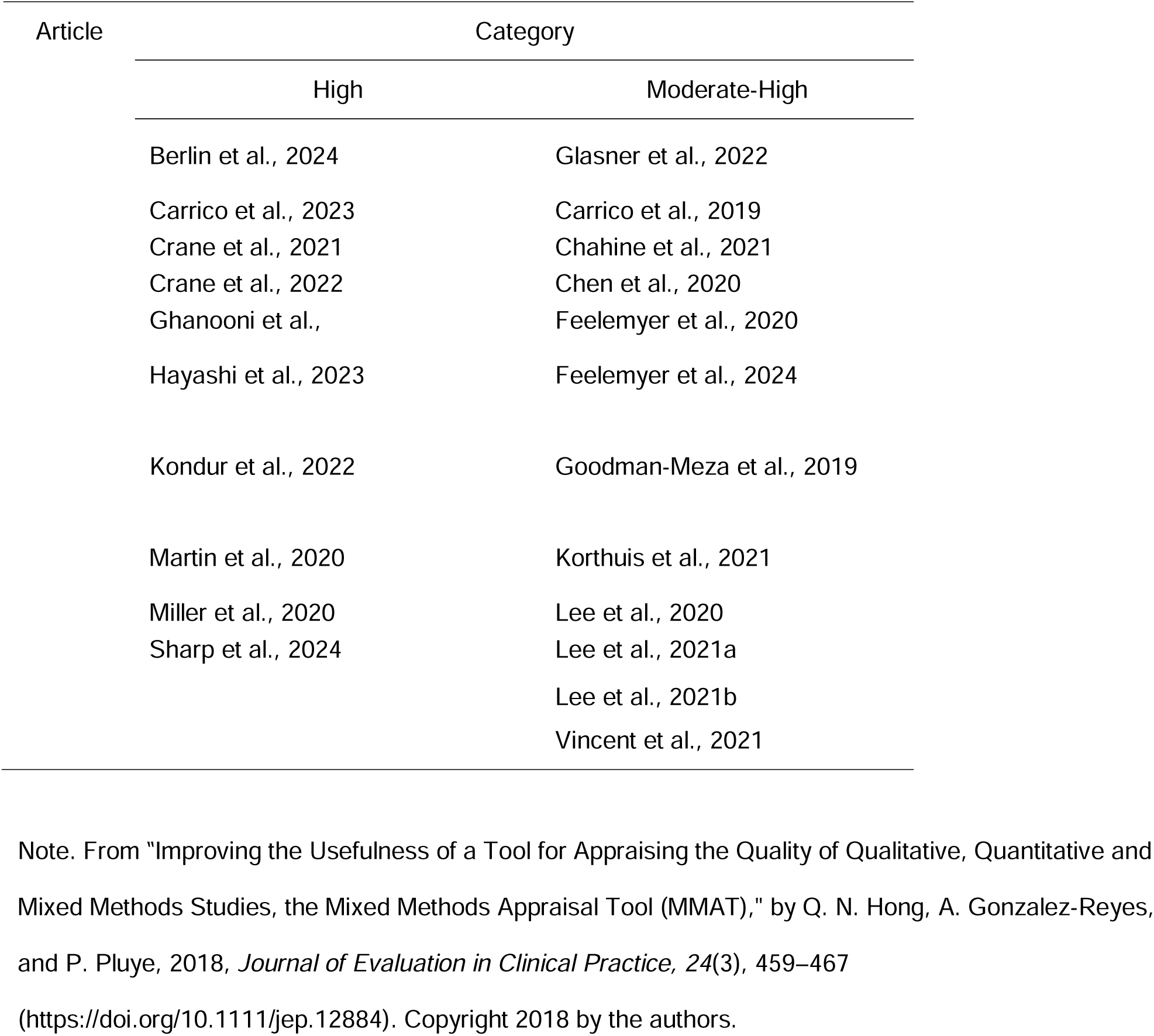
Quality Assessment Category by the Mixed Method Appraisal Tool.

**Table 2.**
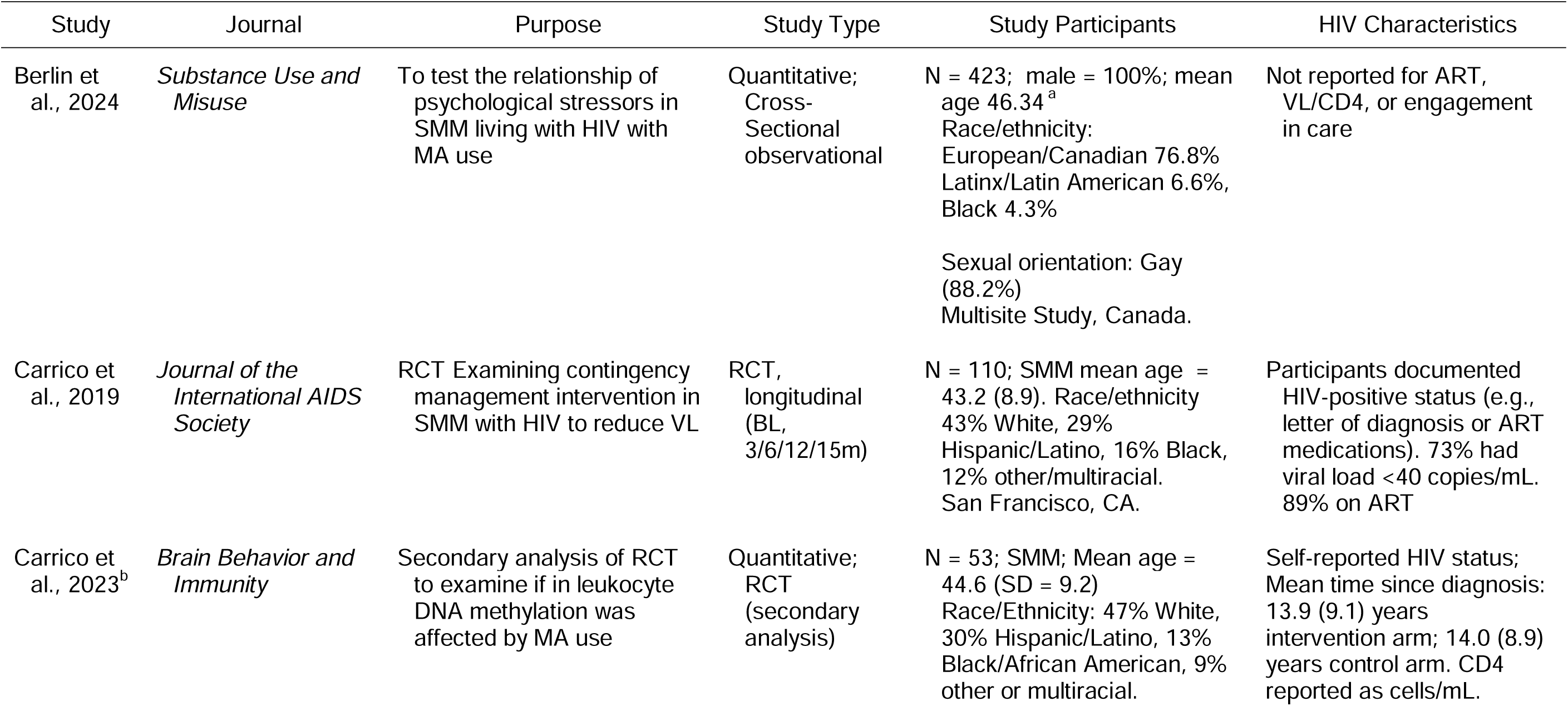
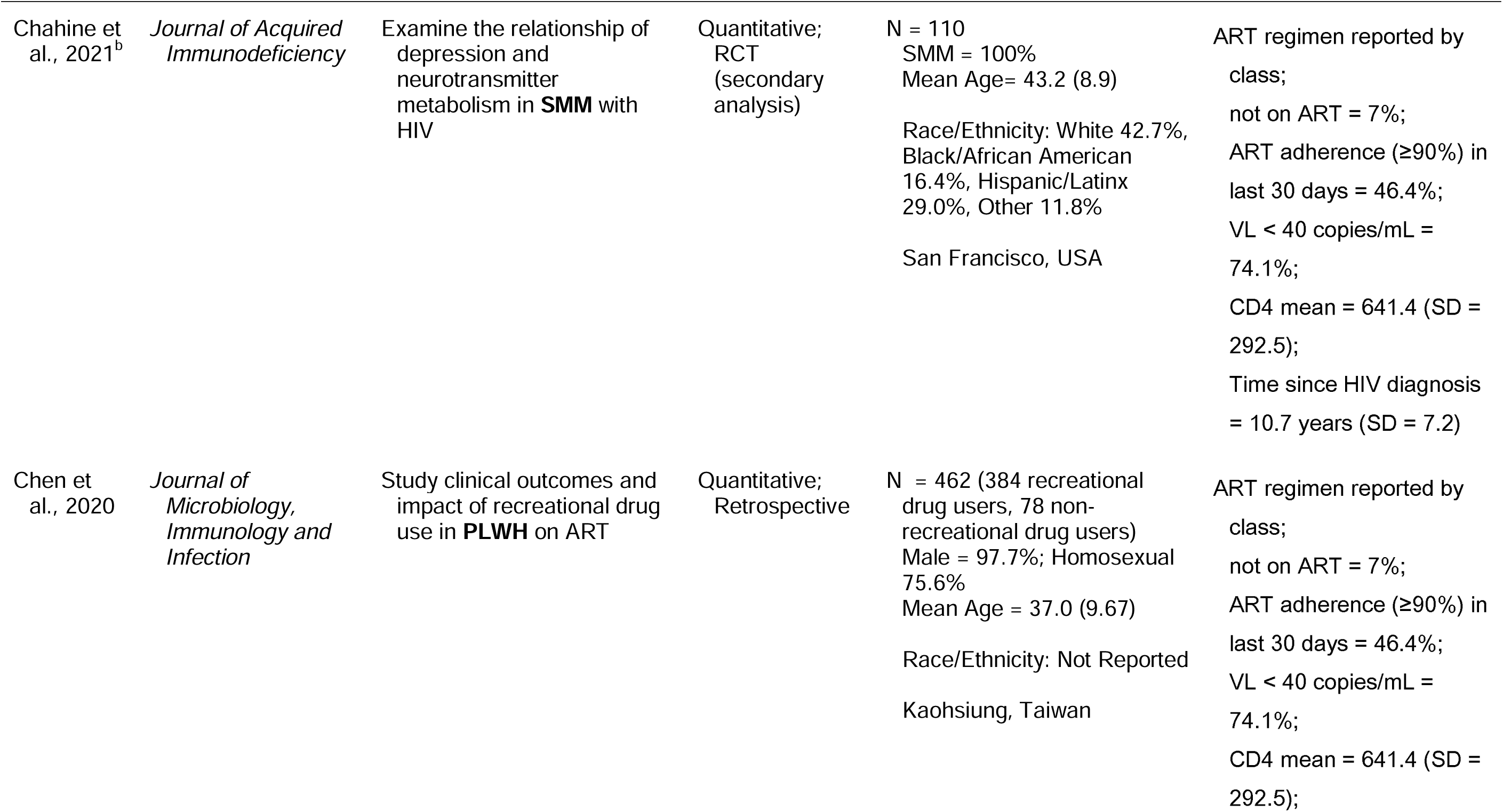
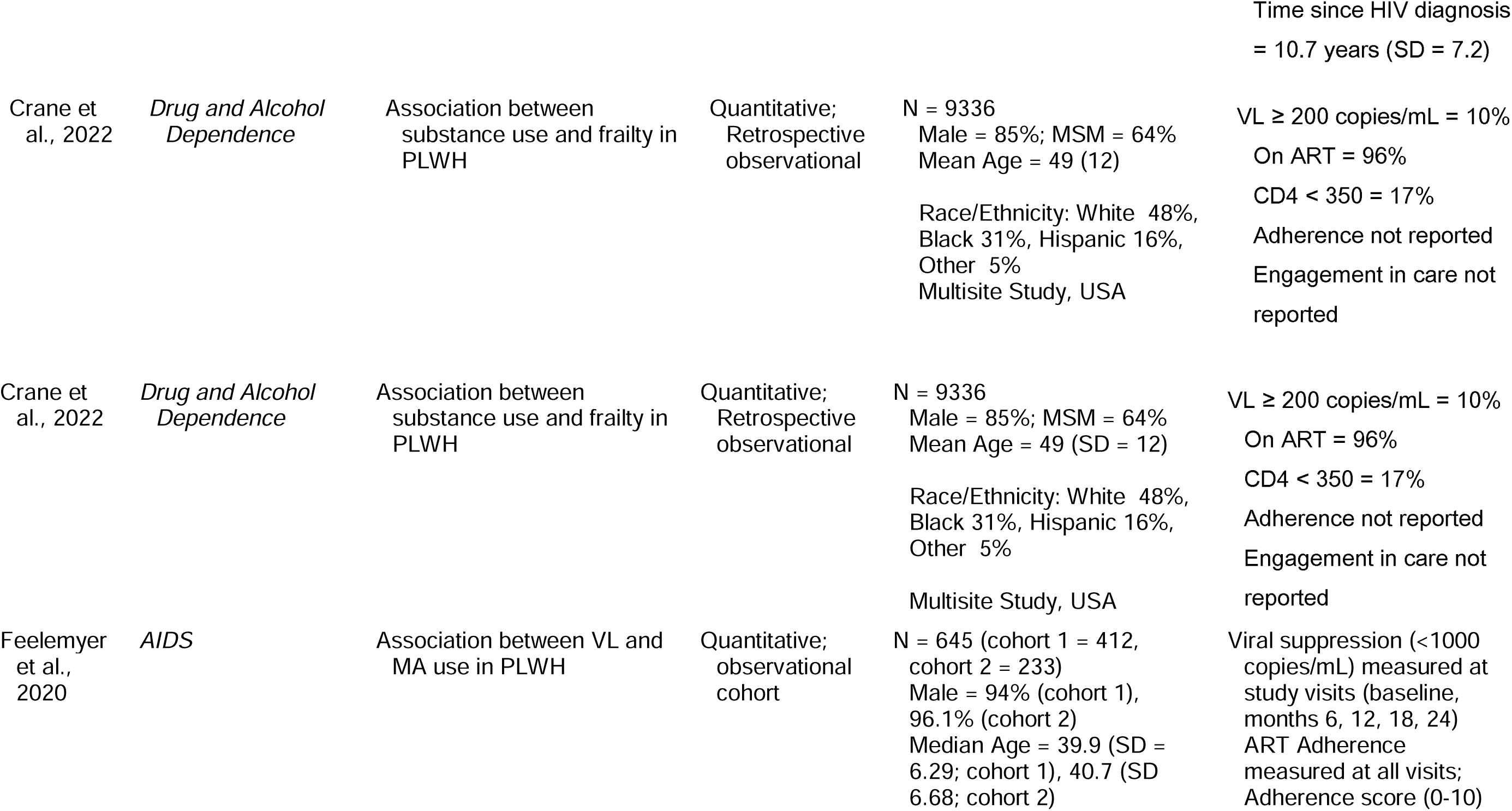
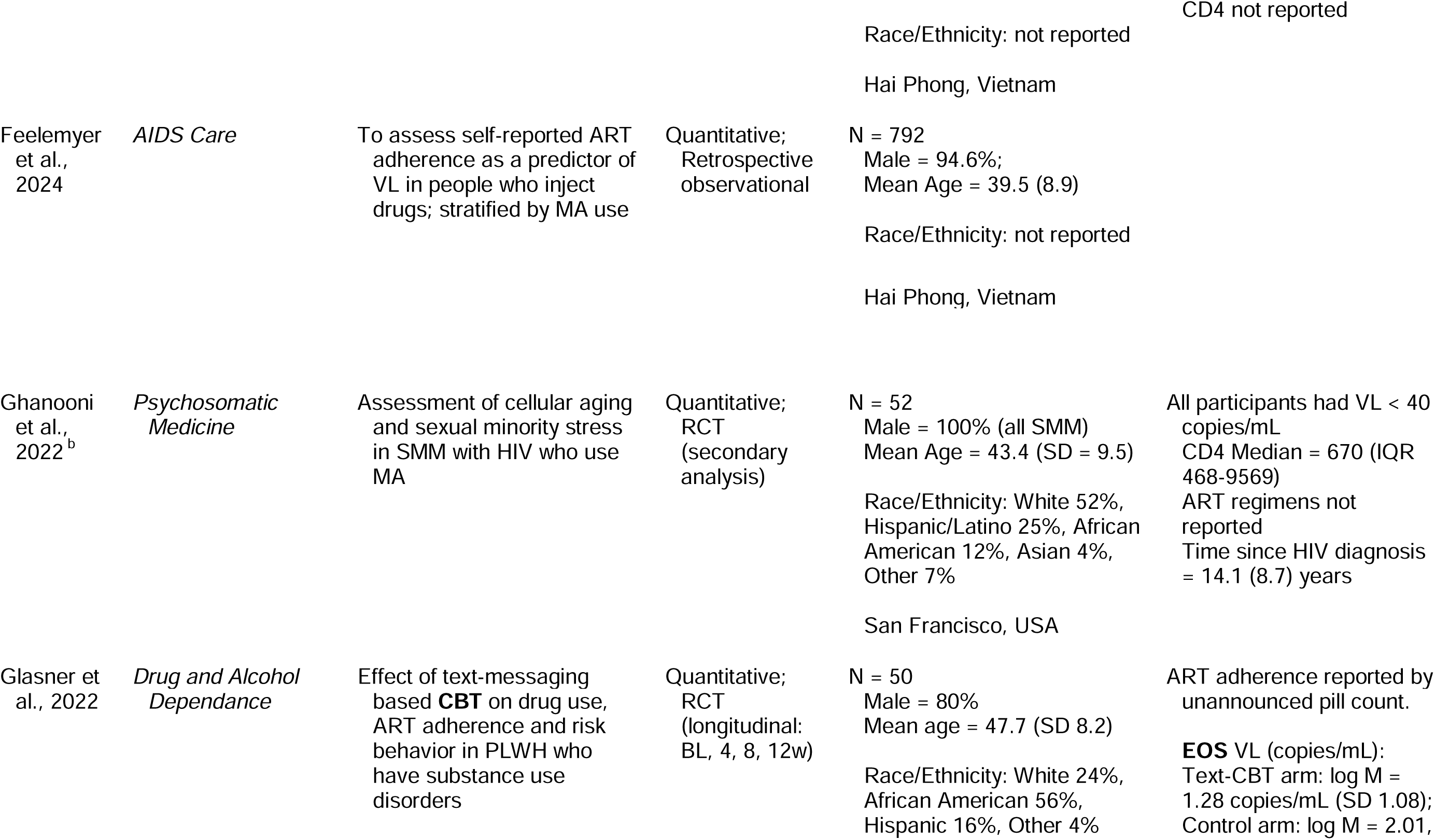
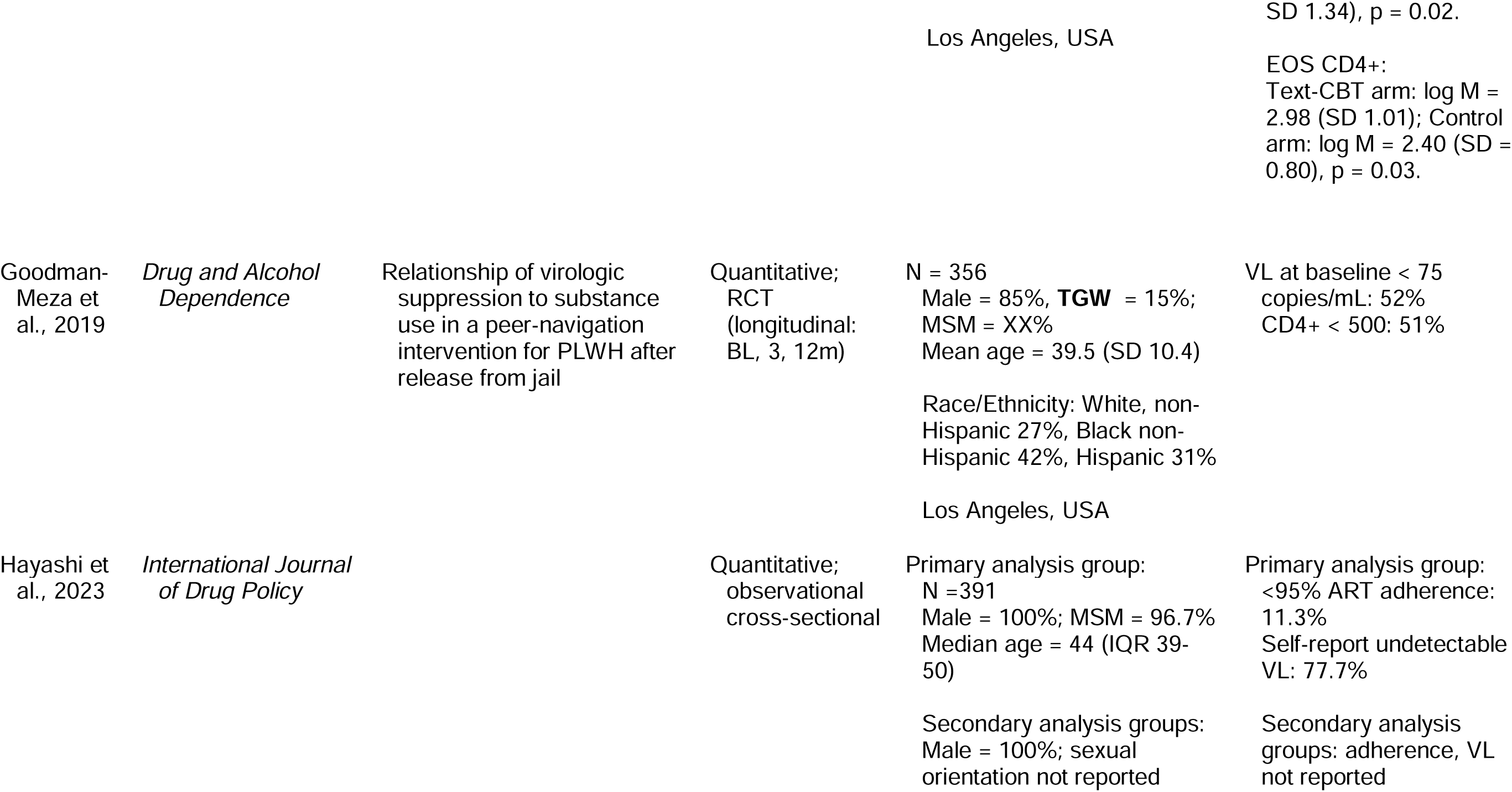
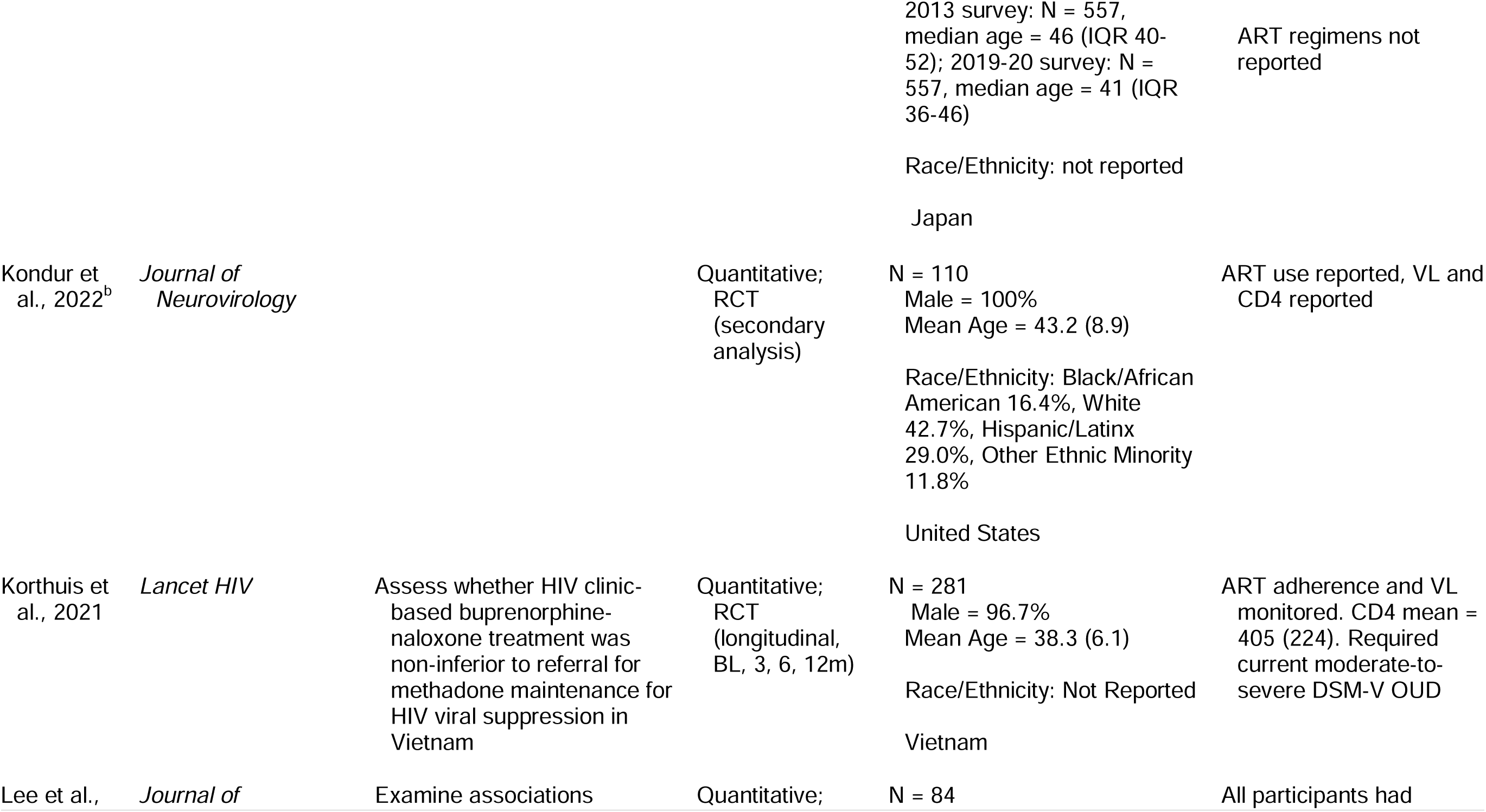
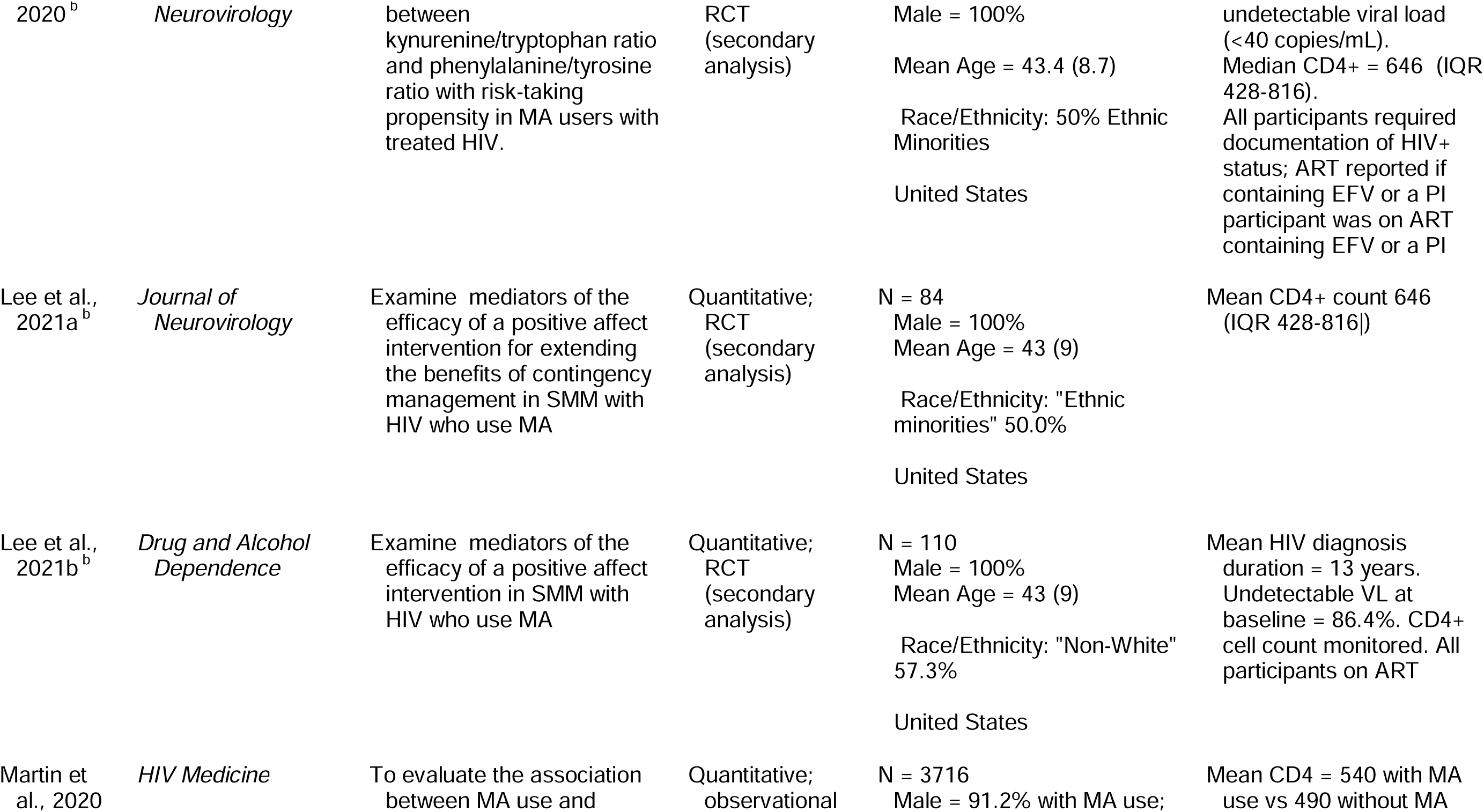
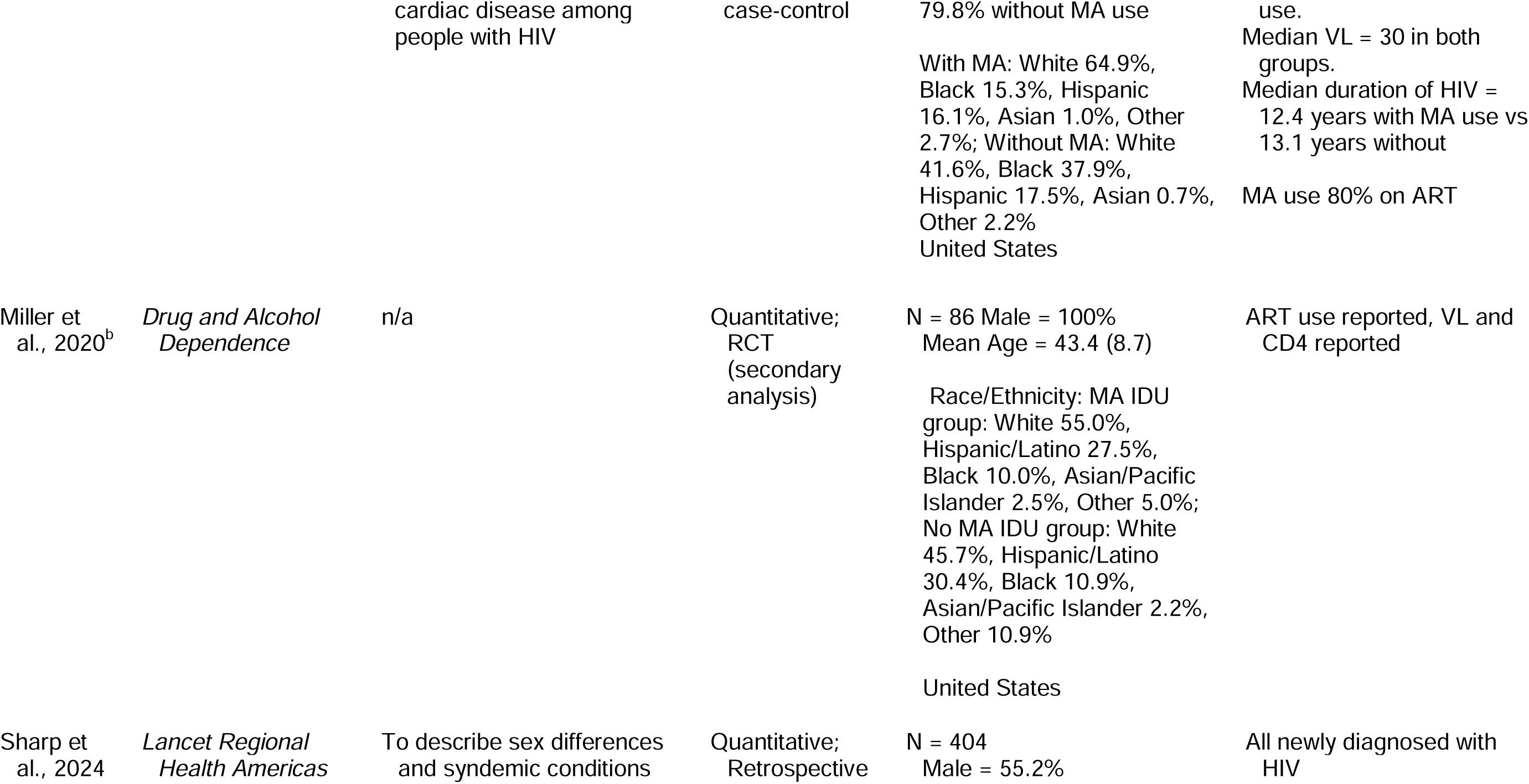
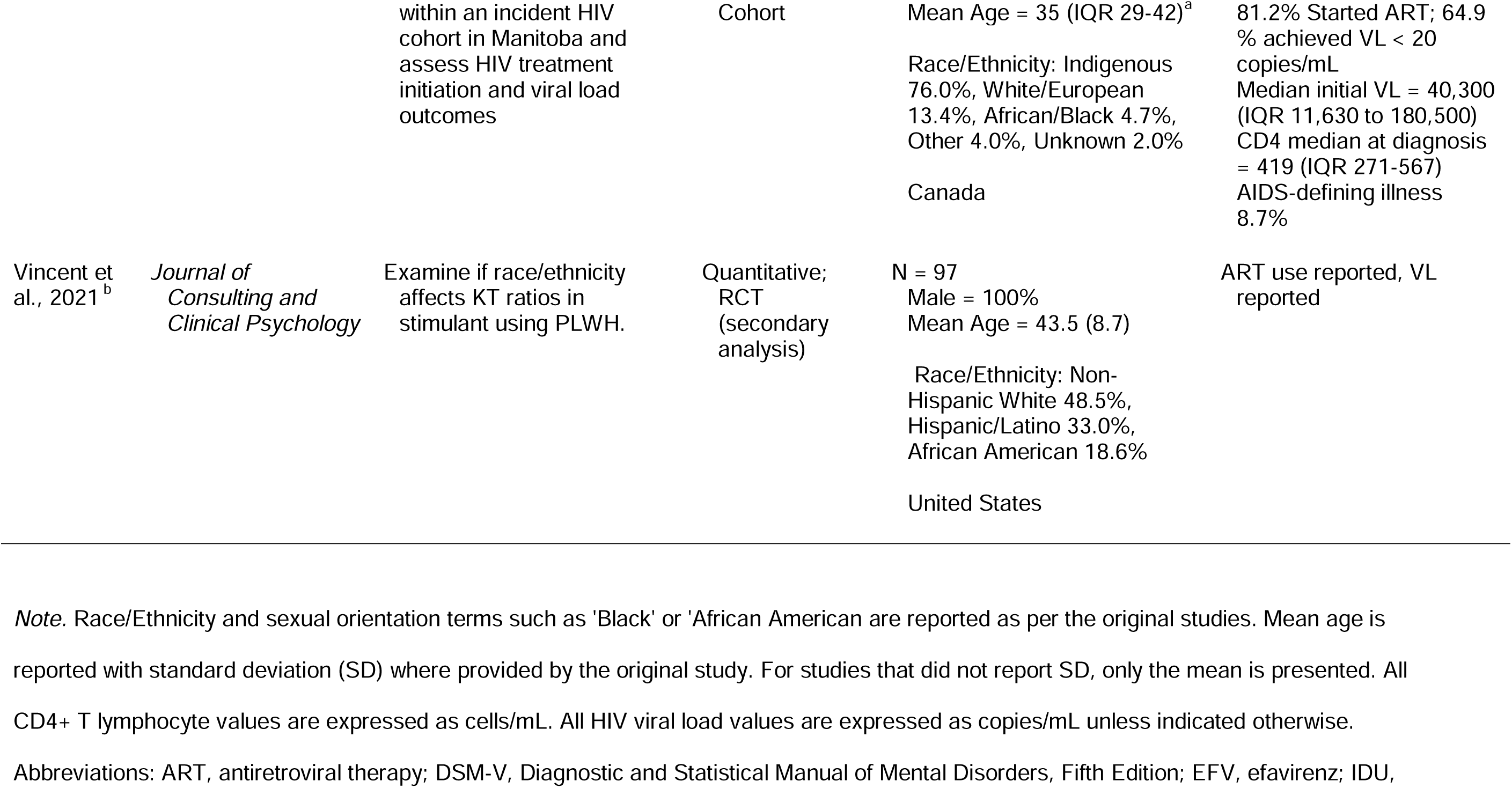
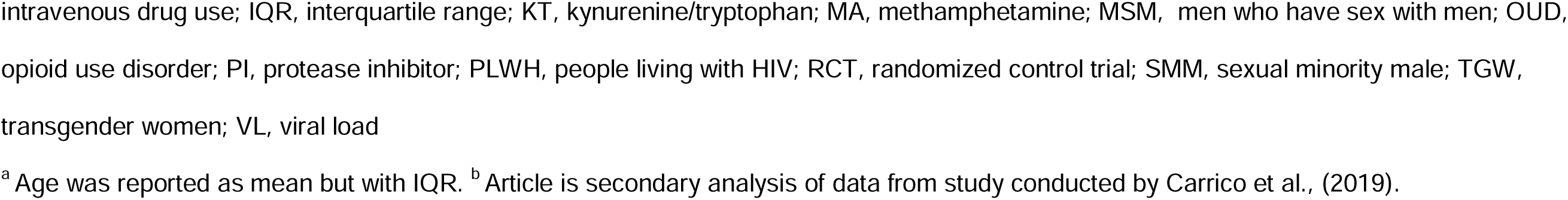
Characteristics of Included Studies.

**Table 3.** Reporting of Methamphetamine Use and Other Substances.

| Study | MA Assessment<br>Self-Report or Questionnaire | MA Assessment<br>Biological Assay | MA Route of<br>Administration | Other Substances Reported |
| --- | --- | --- | --- | --- |
| Berlin et al.,<br>2024 | Self-report: MA use in lifetime,<br>last 6 months, or never | Not assessed | Not assessed | Not assessed |
| Carrico et al.,<br>2019 | Self-report: MA use in last 3<br>months (Y/N); Frequency<br>rated on a Likert-like scale<br>from 0 (not at all) to 7 (daily) | Urine: every study<br>visit; iCup<sup>a</sup>;<br>(positive for any<br>stimulant);<br>Negative urine at<br>BL reflexed to hair | Not assessed | Cocaine, powder; cocaine, crack |
| Carrico et al.,<br>2023<sup>b</sup> | Self-report: MA use in last 3<br>months (Y/N); Frequency<br>rated on a Likert-like scale<br>from 0 (not at all) to 7 (daily) | Urine: every study<br>visit; iCup<sup>a</sup>;<br>(positive for any<br>stimulant);<br>Negative urine at<br>BL reflexed to hair | Not assessed | Cocaine, powder; cocaine, crack |
| Chahine et<br>al., 2021<sup>b</sup> | Self-report: MA use in last 3<br>months (Y/N); Frequency<br>rated on a Likert-like scale<br>from 0 (not at all) to 7 (daily) | Urine: every study<br>visit; iCup<sup>a</sup>;<br>(positive for any<br>stimulant);<br>Negative urine at<br>BL reflexed to hair | Not assessed | Cocaine; prescription stimulants<br>excluded from study |
| Chen et al.,<br>2020 | Self-report: MA use in the past 6<br>months (yes/no)<br>Frequency of use not<br>assessed | Not assessed | Not assessed;<br>IV drug use assessed<br>as separate variable<br>without regard to type<br>(12.0% of recreational<br>drug use arm) | Self-report; use in the past 6 months:<br>nitrates, MDMA, amphetamine,<br>ketamine, marijuana |

**Table 3 (continued)***Reporting of Methamphetamine Use and Other Substances*
| Study | MA Assessment<br>Self-Report or Questionnaire | MA Assessment<br>Biological Assay | MA Route of<br>Administration | Other Substances Reported |
| --- | --- | --- | --- | --- |
| Study | MA Assessment<br>Self-Report or Questionnaire | MA Assessment<br>Biological Assay | MA Route of<br>Administration | Other Substances Reported |
| Crane et al.,<br>2021 | Modified ASSIST: Current MA<br>use in past 3 months (7.5%)<br>Frequency of use not<br>assessed | Not assessed | Not assessed; IV drug<br>use assessed as a<br>separate variable<br>without regard to type<br>(2.7%) | Self-report; use in the past 6 months:<br>marijuana, cocaine/crack, illicit<br>opioid, IDU, ETOH |
| Crane et al.,<br>2022 | Modified ASSIST: Current MA<br>use in past 3 months (15%),<br>former MA use (26%) or<br>never use (59%)<br>Frequency of use not<br>assessed | Not assessed | Not assessed; IV drug<br>use assessed as a HIV<br>transmission risk factor<br>(12%) | Self-report; current/former/never use:<br>any drug use, cocaine/crack,<br>marijuana, illicit opioid, cigarette,<br>e-cigarette, alcohol |
| Feelemyer et<br>al., 2020 | Self-reported use in last 30 days<br>at study visits (baseline,<br>month 6, 12, 18, 24):<br>no MA use, moderate (1-19<br>days use), heavy (20+ days<br>use) | Not assessed | Not assessed | Each study visit:<br>Self-report in last 30 days: Alcohol,<br>street methadone<br>Urine screen for morphine |
| Feelemyer et<br>al., 2024 | Self-reported used in the last 30<br>days: recent use vs. no<br>recent use; number of days<br>of MA use reported by those<br>with recent use (0 days per<br>month: 57.4%, 1-19 days per<br>month: 34.5%, 20-30 days<br>per month: 8.3%) | Not assessed | Not assessed;<br>Participants were self-reported injecting drug<br>users but did not<br>specify type of drug | Self-report: number of days injecting<br>heroin in past 30 days; current<br>(yes/no): methadone, cannabis,<br>alcohol, binge drinking |

**Table 3 (continued)***Reporting of Methamphetamine Use and Other Substances*
| Study | MA Assessment<br>Self-Report or Questionnaire | MA Assessment<br>Biological Assay | MA Route of<br>Administration | Other Substances Reported |
| --- | --- | --- | --- | --- |
| Ghanooni et al., 2022<sup>b</sup> | Self-report: MA use in last 3 months (Y/N); Frequency rated on a Likert-like scale from 0 (not at all) to 7 (daily) | Urine: every study visit; iCup; hair | Not assessed | Cocaine |
| Glasner et al., 2022 | Modified TLFB structured interview asked about drug use in prior 90 days (baseline) or 30 days (4-, 8-, 12- week visits).<br>MA use summary statistics not reported in article | Urine: every study visit (CLIAwaved, Inc). | Not assessed; BRA recorded injection drug use and non-injection drug use in previous 30 days but did not specify type of drug | Self-report: substance use previous using TLFB (previous 90 days at baseline, previous 30 days at subsequent visits).<br>Urine screening detected amphetamines, benzodiazepines, methadone, cocaine, methamphetamine, barbiturates, oxycontin, opioids, marijuana |
| Goodman-Meza et al., 2019 | Standardized questionnaire with questions about methamphetamine or amphetamines<br>Baseline: use in 30 days prior to arrest; follow-up visits: use in past 30 days<br>Duration of use measured in years<br>MA use 58% | Not assessed | Not assessed; PWID listed as risk factor (mutually exclusive with MSM, TWG and none of the above), 15% at baseline | Self-reported use of Cocaine, heroin, prescription opioids, binge alcohol<br>Baseline: use in 30 days prior to arrest; follow-up visits: use in past 30 days<br>High-risk drug use reported separately |

**Table 3 (continued)***Reporting of Methamphetamine Use and Other Substances*
| Study | MA Assessment<br>Self-Report or Questionnaire | MA Assessment<br>Biological Assay | MA Route of<br>Administration | Other Substances Reported |
| --- | --- | --- | --- | --- |
| Hayashi et al., 2023 | Primary analysis group: MA use (yes/no): ever (37.1%), past year (10.2%)<br>Secondary analysis group: MA use (yes/no) in past year: 2019-2020 group (6.6%), 2013 group (4.3%) | Not assessed | Not assessed; primary analysis group reported any IDU in past year (10.0%) and ever (35.5%) | Primary analysis group: Self-reported use: ever and in the past year; Secondary analysis group: self-reported use in the past year only of: alkyl nitrates, 5-MeO-DPIT, NPS<sup>c</sup>, MDMA, cannabis, air duster |
| Kondur et al., 2022<sup>b</sup> | Yes, Lifetime use | Urine: every study visit; iCup<sup>a</sup>; (positive for any stimulant); Negative urine at BL reflexed to hair | Not assessed | Other stimulant, ETOH |
| Korthuis et al., 2021 | Not Reported | Urine testing at baseline, then every 3 months through 12 months | Not assessed | Other stimulant, Heroin, Methadone/Other opioid substitution therapy |
| Lee et al., 2020<sup>b</sup> | Not Reported | Urine: every study visit; iCup<sup>a</sup>; (positive for any stimulant); Negative urine at BL reflexed to hair | Not assessed | Cocaine - Any |

**Table 3 (continued)***Reporting of Methamphetamine Use and Other Substances*
| Study | MA Assessment<br>Self-Report or Questionnaire | MA Assessment<br>Biological Assay | MA Route of<br>Administration | Other Substances Reported |
| --- | --- | --- | --- | --- |
| Lee et al.,<br>2021a<sup>b</sup> | Not Reported | Urine: every study visit; iCup<sup>a</sup>; (positive for any stimulant); Negative urine at BL reflexed to hair | Not assessed | Cocaine detected through biological assay |
| Lee et al.,<br>2021a<sup>b</sup> | Number of days using stimulants in past 30 days assessed at baseline, 3, 6, 12, and 15 months | Urine: every study visit; iCup; (positive for any stimulant); Negative urine at BL reflexed to hair | Not assessed | Urine screening detected any MA or cocaine use; prescription stimulant use excluded from study |
| Martin et al.,<br>2020 | Methamphetamine abuse/dependence based on DSM-IV criteria Both lifetime and current (past 30 days) | Not assessed | Not assessed | Cocaine, Alcohol |
| Miller et al.,<br>2020<sup>b</sup> | Not Reported | Urine: iCup<sup>a</sup>; (positive for any stimulant); Negative urine reflexed to hair | Injection | Cocaine |
| Sharp et al.,<br>2024 | Not Reported | Not assessed | Self-reported: injection, intranasal, inhaled, smoked | Cocaine - Any, Marijuana/Cannabis, Opioid (Any) |

**Table 3 (continued)***Reporting of Methamphetamine Use and Other Substances*
| Study | MA Assessment<br>Self-Report or Questionnaire | MA Assessment<br>Biological Assay | MA Route of<br>Administration | Other Substances Reported |
| --- | --- | --- | --- | --- |
| Vincent et al.,<br>2021<sup>b</sup> | Not Reported | Urine: every study<br>visit; iCup<sup>a</sup>;<br>(positive for any<br>stimulant);<br>Negative urine at<br>BL reflexed to hair | Not assessed | Cocaine - Any, Other stimulant |

Note. Abbreviations: ART, antiretroviral therapy; BRA, behavioral risk assessment (Reback, 2005); DPT, N,N-Dipropyltryptamine; DSM-IV, Diagnostic and Statistical Manual of Mental Disorders, Fourth Edition; ETOH, alcohol; IDU, intravenous drug use; IQR, interquartile range, MA, methamphetamine; MDMA, 3,4-Methylenedioxymethamphetamine; MSM, men who have sex with men; NPS, new psychoactive substances; OUD, opioid use disorder; PI, protease inhibitor; PLWH, people living with HIV; PWID, people who inject drugs; RCT, randomized control trial; SMM, sexual minority male; TGW, transgender women; TLFB, timeline follow back; VL, viral load
<sup>a</sup>iCup urine screen, Redwood Biotech Inc., Santa Rosa, Ca. <sup>b</sup>Article is secondary analysis of data from study conducted by Carrico et al., (2019). <sup>c</sup>NPS defined as “Includes variations of synthetic cannabinoids, synthetic cathinones, phenethylamine, thiophene derivatives, etc.” (Hayashi et al.)

### Demographic Information

The study populations’ gender was disproportionately male, with 12 studies comprised of exclusively male participants(Berlin et al., 2024; Carrico et al., 2019, 2024; Chahine et al., 2021; Ghanooni et al., 2022; Hayashi et al., 2023; Kondur et al., 2022; J. Lee et al., 2020; J.-Y. Lee, Glynn, et al., 2021; J.-Y. Lee, Lee, et al., 2021; Miller et al., 2020; Vincent et al., 2021). Among these male-only studies, all except one (Berlin et al., 2024) focused specifically on men who have sex with men (MSM) or sexual minority men (SMM). Gender composition varied in studies that did include a female population, ranging from 2.3% to 44.5%. Gender identity was explicitly reported in only one study (Goodman-Meza et al., 2019), with 15% of the study population transgender female. Across the nine studies that reported race/ethnicity data (N=22,134), the weighted mean percentages showed that participants were predominantly White/European (44.7%, range: 13.4%-76.8%), followed by Black/African American (33.2%, range: 4.3%- 56.0%), and Hispanic/Latino (15.4%, range: 6.6%-31.0%). A smaller proportion were categorized as Other/Multiracial (4.9%, range: 2.4%-11.8%). Notably, one study from Canada (Sharp et al., 2024) reported a large Indigenous population (76.0% of N=404). The five Asian studies did not report race or ethnicity data (Chen et al., 2021; Feelemyer et al., 2020, 2024; Hayashi et al., 2023; Korthuis et al., 2021). Three studies did not provide specific race or ethnicity but instead reported the total percentage of “ethnic minorities” (Lee et al., 2020), “person of color” (Lee et al., 2021b), or “non-White” (Lee et al., 2021a). However, these studies were all secondary analyses of the Carrico et al. (2019) primary article which did report race and ethnicity data. The studies that used the Carrico et al. (2019) article as the primary data source were excluded from the demographic analysis to avoid counting them twice.

### HIV-Related Data

For confirmation and verification of HIV status, most studies used multiple methods. Only one relied strictly on self-reporting (Berlin et al., 2024). Nine studies involved participants who were already patients at an HIV clinic or on an HIV clinical trial without further information (Chen et al., 2021; Feelemyer et al., 2020; Goodman-Meza et al., 2019; Hayashi et al., 2023; Korthuis et al., 2021; Lee et al., 2020b; Martin et al., 2020; Miller et al., 2020). Four studies explicitly stated review of medical records (Crane et al., 2021; Crane et al., 2022; Kondur et al., 2022), with three additional studies requiring a medication bottle or prescription for ART (Carrico et al., 2023; Lee et al., 2020; Lee et al., 2020a). Four studies required baseline laboratory testing, including HIV point-of-care testing (Feelemyer et al., 2020; Feelemyer et al., 2024; Ghanooni et al., 2022; Sharp et al., 2024). The reporting of key HIV clinical parameters— viral load (VL), CD4 count, and antiretroviral therapy (ART) use—varied across studies. Most studies (19 of 22) reported whether participants were receiving ART. Viral load and CD4 measurements were reported in varying combinations: fourteen studies reported both VL and CD4 counts (Carrico et al., 2023; Chahine et al., 2021; Chen et al., 2021; Crane et al., 2021; Crane et al., 2022; Ghanooni et al., 2022; Glasner et al., 2022; Goodman-Meza et al., 2019; Kondur et al., 2022; Lee et al., 2020; Lee et al., 2021a; Lee et al., 2021b; Martin et al., 2020; Miller et al., 2020), while four studies reported VL only (Feelemyer et al., 2020; Feelemyer et al., 2024; Hayashi et al., 2023; Vincent et al., 2021), and one study reported CD4 only (Korthuis et al., 2021). Three studies did not report VL or CD4 measurements (Berlin et al., 2024; Carrico et al., 2019; Sharp et al., 2024). The frequency of these measurements varied by study design, with longitudinal studies and RCTs typically including regular monitoring at specified intervals, while cross-sectional studies reported single time-point measurements.

### Methamphetamine Use

Methamphetamine use was measured using biologic and self-reported measures. Studies reported urine screening (Chahine et al., 2021; Feelemyer et al., 2020; Feelemyer et al., 2024; Ghanooni et al., 2022; Glasner et al., 2022; Korthuis et al., 2021; Lee et al., 2020b) or a combination of hair or urine screening (Carrico et al., 2023; Carrico et al., 2019; Kondur et al., 2022; Lee et al., 2020; Lee et al., 2020a; Miller et al., 2020; Vincent et al., 2021). No studies reported using serum assays for screening. Six studies reported using an iCup urine test for MA use (Redwood Biotech Inc., Santa Rosa, Ca) (Ghanooni et al., 2022; Kondur et al., 2022; Lee et al., 2020; Lee et al., 2020a; Lee et al., 2020b; Miller et al., 2020), a lateral flow assay which reports stimulant use in the past 72 hours. Of these six studies, all except for one (Lee et al., 2020b) explicitly aimed to study MA use.

Few studies reported details regarding the route of MA administration. Only four articles mentioned the route of administration (Feelemyer et al., 2020; Glasner et al., 2022; Miller et al., 2020; Sharp et al., 2024); all others only captured MA use as yes/no. Even in studies that discussed the route of MA use, the specifics were unclear. Miller et al. (2020) examined substance use in the context of a CBT intervention; MA use was self-reported during structured interviews, and injection use was recorded as a drug-related risk behavior, but there is no discussion of the overlap or exclusivity of these criteria. Feelemyer et al. (2020) examined MA use in persons who inject drugs in Vietnam but did not capture if non-injection MA use was occurring. They report that 100% of the population was injecting heroin at baseline, and MA use was captured by self-report, but the route of administration was not discussed. Glasner et al. (2022) assessed injection drug use independently from MA use without discussion of overlap or exclusivity. No studies discussed snorting or rectal administration of MA. The context of MA use was frequently assessed alongside other substance use. Most studies reported concurrent use of other substances, particularly cocaine, cannabis, and alcohol. However, the methodology for assessing polysubstance use varied widely, from comprehensive toxicology panels to single-item self-report measures. The temporal reporting of MA use also varied. Studies captured different periods, including lifetime use, past year use, past six months, past three months, past month, and current use. For example, Glasner et al. (2022) assessed use in the past three months, while Berlin et al. (2024) focused on the past six months. The variation in time frames makes direct comparisons between studies challenging and highlights the need for standardized reporting periods in future research.

### Strengths and Weaknesses

This systematic review represents the first comprehensive assessment of methodologies used to report MA use among people living with HIV. The review has several notable strengths, including its broad interdisciplinary scope spanning HIV research, virology, neurology, and addiction science. By identifying studies that captured MA use through both biometric and psychosocial measures, this review provides a comprehensive representation of current measurement approaches in the literature. The included studies demonstrated considerable ethnic diversity in those that reported race and ethnicity data.

Several important limitations in this review must be acknowledged. Of the 22 studies identified, five were secondary analyses of data collected by Carrico et al. (2019), and nine articles shared lead authors. While these can present different data from the primary studies, this overlap suggests the true diversity of evidence may be more limited than initially identified. The absence of qualitative or mixed-methods studies represents another significant limitation, as these approaches could provide valuable experiential insights into MA use in this population with the ability to capture contextual nuance. Geographic representation was limited to North America and Asia, with no studies from Africa, Europe, or Latin America, restricting the generalizability of findings. Additionally, study populations were heavily male, with notable underrepresentation of women and limited attention to gender identity. This is particularly concerning given evidence suggesting higher rates of substance use among transgender individuals compared to cisgender population (Cotaina et al., 2022; Ruppert et al., 2021).

A fundamental challenge identified across studies was the inconsistent classification and measurement of MA use. Many studies reported on broader categories, such as "stimulant use" or "injection drug use," which could include substances beyond MA. When biological assays were employed, they predominantly relied on urine screening using the iCup test, which does not distinguish between MA and cocaine. While self-reported MA use was common, many studies’ lack of biological validation could have added valuable context to and strengthened the data.

### Implications

Future studies should prioritize establishing standardized definitions and measurement approaches for MA use that clearly differentiate it from other substances. While biological assays could be a useful tool to help validate self-reporting of MA, their implementation must also consider their costs and the limited detection windows. More comprehensive reporting of MA administration routes (smoking, injecting, snorting) can identify if they have an impact on HIV-related outcomes and comorbidities in this population. Expanding geographic and demographic representation, particularly regarding gender identity and women, should be prioritized.

These findings have implications for clinical practice. The current lack of standardization in measuring and documenting MA use challenges healthcare providers. While biological assays offer objective evidence of recent use, cost constraints can make them impractical for routine clinical assessment. Patient self-disclosure is essential but can be subject to social desirability bias and stigma-related underreporting. The lack of specificity in current ICD-10 diagnosis codes for distinguishing between different stimulant use disorders further complicates clinical documentation and continuity of care.

## Conclusion

This review highlights significant variability in how MA use is solicited from PLWH and documented across various types of research. There are still many unanswered questions about the use of MA in the setting of HIV. Identification of methodologies in reporting can help develop a future picture of the impact on this population and guide further research and clinical practices.

## Data Availability

All data produced in the present work are contained in the manuscript

### Appendix A

#### Sample Search Terms for EMBASE

1. ’human immunodeficiency virus’/exp
2. ’HIV’/exp
3. ("HIV" OR "human immunodeficiency virus").tw,kw
4. ’methamphetamine’/exp
5. ’stimulant’/exp
6. ("methamphetamine" OR "stimulant*" OR "crystal meth").tw,kw
7. ’screening’/exp
8. ’questionnaire’/exp
9. ’data collection method’/exp
10. ("reporting" OR "screening" OR "questionnaire" OR "data collection").tw,kw
11. "preexposure prophylaxis".ab,kw,ti OR "pre-exposure prophylaxis".ab,kw,ti OR "prep".ab,kw,ti
12. ("human immunodeficiency virus infected patient").ab,kw,ti
13. "injection drug user".tw,kw
14. (’preexposure prophylaxis’ OR ’prep’).ab,kw,ti
15. ("methamphetamine" OR "stimulant").ab,kw,ti
16. ("reporting" OR "screening" OR "questionnaire" OR "data collection").ab,kw,ti
17. ("preexposure prophylaxis" OR "prep").ab,kw,ti
18. ("human immunodeficiency virus infection").exp
19. ("middle-aged" OR "adult" OR "aged" OR "young adult" OR "very elderly").lim
20. (2019 OR 2020 OR 2021 OR 2022 OR 2023 OR 2024 ).lim

### Appendix B

#### Codebook Items for Article Extraction

*Appendix B*

#### Code Book Items

##### Article Information

Author(s):

Title:

Year:

Journal:

Study Type: Qualitative, Quantitative, Mixed-Method

Study Design:

Study Population Size:

Sampling Method:

Population Age Range:

Population Gender:

Population Sexual Orientation:

##### HIV Characteristics

HIV Diagnosis Method: Self-Report, Biological Assay, Medical Record

Reported if in Care:

Reported if on ART: (if yes, are classes of ART listed?)

Reported VL?

Reported CD4?

##### Methamphetamine Use

Method of Capturing Use:

Self-Report? Yes/No

If Yes:
Participant or Clinician/Study Staff Administered?
Is Screening Instrument Listed?

Biological Testing: Yes/No

If Yes:
Type of Assay: Urine/Hair/Serum
Frequency of Testing: Baseline Yes/No, Follow-up testing?

Medical Record Review/Diagnosis Code?

If Yes:
Chart Extraction: Yes/No
Diagnosis Codes: Yes/No
Are diagnosis codes (ICD-10) given?

Qualification of Use:

Current Methamphetamine Use?

Last Methamphetamine Use?

If Yes: Time of last use?

Frequency of use Reported? Yes/No

If Yes: What are frequency options?

Route of Use:

Captured? Yes/No

If Yes: What options are listed

Contextual Factors Related to Use? Yes/No

If Yes:

Social/Behavioral Factors?

Other Substance Use Recorded? Yes/No

If Yes:

Specify (i.e. ETOH, cocaine, opioids, marijuana)

Other Comorbidities Reported? Yes/No

If Yes:

Specify (i.e. syphilis, gonorrhea, hepatitis B, hepatitis C, HPV)

**Findings:**

**Recommendations:**

**Strengths:**

**Weakness**

## Notes

### Competing Interest Statement

The authors have declared no competing interest.

### Funding Statement

This study did not receive any funding

